# Development and Presentation of an Objective Risk Stratification Tool for healthcare workers when dealing with the COVID-19 pandemic in the UK: Risk modelling based on hospitalisation and mortality statistics compared to epidemiological data

**DOI:** 10.1101/2020.05.05.20091967

**Authors:** W David Strain, Janusz Jankowski, Angharad Davies, Peter MB English, Ellis Friedman, Helena McKeown, Su Sethi, Mala Rao

**Author notes:** Corresponding Author:* Dr David Strain, Diabetes and Vascular Medicine Research, University of Exeter, College of Medicine and Health, Royal Devon & Exeter Hospital, Barrack Road, Exeter, EX2 5AX. Declarations of Interest; None to declare by all authors.

## Abstract

**Objectives:** Healthcare workers have a greater exposure to individuals with confirmed SARS-novel coronavirus 2, and an estimated 5-fold higher probability of contracting coronavirus disease (COVID)-19, than the general population. Many organisations have called for risk assessments to be put in place to minimise this risk. We wished to explore the predictive role of basic demographics in order to establish a simple tool that could help risk stratify healthcare workers.

**Setting:** We undertook a review of the published literature (including multiple search strategies in MEDLINE with PubMed interface) and critically assessed early reports on medRxiv, a pre-print server (https://www.medrxiv.org: date of last search: December 21, 2020). We explored the relative risk of mortality from readily available demographics in order to identify the population at highest risk.

**Results:** The only published studies specifically assessing the risk of healthcare workers had limited demographics available, therefore we explored the general population in the literature.

*Clinician Demographics:* Mortality increased with increasing age from 50 years onwards. Male sex at birth, people of black and minority ethnicity groups had higher susceptibility to both hospitalisation and mortality. *Co-morbid Disease*. Vascular disease, renal disease, diabetes and chronic pulmonary disease further increased risk.

*Risk stratification tool:* A risk stratification tool was compiled using a Caucasian female <50years with no comorbidities as a reference. A point allocated to risk factors associated with an approximate doubling in risk. This tool provides numerical support for healthcare workers when determining which team members should be allocated to patient facing clinical duties compared to remote supportive roles.

**Conclusions:** We have generated a tool which can provide a framework for objective risk stratification of doctors and health care professionals during the COVID-19 pandemic, without requiring disclosure of information that an individual may not wish to share with their direct line manager during the risk assessment process.

**Strengths and limitations of this study:**

- There is an increased risk of mortality in the clinical workforce due to the effects of COVID-19.
- This manuscript outlines a simple risk stratification tool that helps to quantify an individual’s biological risk
- This will assist team leaders when allocating roles within clinical departments.
- This tool does not incorporate other external factors, such as high-risk household members or those at higher risk of mental health issues, that may require additional consideration when allocating clinical duties in an appropriate clinical domain.
- This population-based analysis did not explain for the very high risk observed in BAME healthcare workers suggesting there are other issues at play that require addressing.

---

The Health and Safety executive mandate that all employers protect their employees from harm under the Management of Health and Safety at Work Regulations 1999. There are three key elements to this, identify what could cause the injury (the hazard), decide how likely that someone could be harmed and how seriously they are likely to be harmed (the vulnerability), and what actions can be taken to minimise this risk (the mitigation). In the current coronavirus pandemic, it is clear the coronavirus disease (COVID-19) is the agent that causes injury. The risk of harm is higher in healthcare workers compared to the general population^1^, and thus action is required to minimise this risk. In the early phase of the pandemic, the Office of National Statistics reported medical practitioners had a 2.5-fold increase (95% CI 1.5-4.3) in mortality compared to the average mortality from 2014-2018.^2^. This compared to a 50% increased risk in the age-matched general population (HR 1.5, 95%CI 1.5-1.6). This trend was in keeping with observations in other countries of higher mortality amongst health care workers^3-7^.

Whilst reasonable measures must be taken to protect all staff members from infection, individuals thought to be at particularly vulnerable from infection may require modification of their practice. The Faculty of Occupational Medicine and NHS Employers in England produced recommendations that all health care practitioners should receive an occupational risk assessment^8 9^. These frameworks were borne of the observation that certain ethnic groups appeared to be at higher risk than others^9^, whilst recognising there are several other biological parameters, such as age, male sex, prior cardiovascular disease, and diabetes that were also associated with adverse outcomes. These predictors of hospitalisation, progression to intensive care units, and ultimately death were been reaffirmed in the Public Health England document^10^.

Despite the intention to improve risk assessment in healthcare settings, these frameworks failed to produce an objective tool in order to improve stratification across the health care system. The need for an such a tool is highlighted by the disproportionate impact of COVID-19 on healthcare workers of black, Asian and minority ethnicity (BAME) descent. Up to the 21^st^ April 2020, 36% and 27% of the fatalities came from people of Indian Asian heritage and Black African descent respectively, despite those populations only representing 10% and 6% of the work force. Existing data suggest biological parameters do not account for all this increased risk, raising the possibility of cultural differences in self-assessment of risk or systemic challenges in modification of hazards for people of different ethnic background. Indeed, these cultural challenges have been proposed as a contributor to the increased risk in BAME populations.

Using published data on the demographics of those who have been hospitalised, and ultimately died, due to COVID-19 compared to the general population prevalence in these determinants we have developed an objective risk stratification tool. Creation of such an objective tool that can be applied equally and without favour to all health care practitioners allows biological risk to be evaluated and used to reduce hazard.

## Methods

We reviewed the published literature (including multiple search strategies in MEDLINE with PubMed interface), EMBASE and critically assessed early reports on medRxiv, a pre-print server (https://www.medrxiv.org/) (date of last search: December 21, 2020).

### Eligibility criteria

Studies were included according to the following criteria

#### Search terms

COVID-19, Coronavirus, SARS-CoV2, Coronavirus AND mortality, hospitalisation

#### Participants

As it had already been observed that there were differences in the impact of COVID-19 in different geographic locations and different socio-economic circumstances, we limited the search to reports from the UK.

#### Outcomes

Given there are selection biases in testing for coronavirus, COVID-19 care and reporting, we explored predominantly the ‘hard outcomes’ of admission to the intensive care unit and mortality. Whereas, the occurrence of mild symptoms and asymptomatic disease may have an impact on the health systems ability to function and nosocomial spread, it would not cause significant long term consequences and thus was not considered as an outcome.

### Information sources and search strategy

We searched the following electronic databases: MEDLINE, EMBASE, and the preprint server MedRxiv from inception to 22^nd^ December 2020. Only English language manuscripts were included. The reference lists of included reports were also searched for additional reports. The majority of the existing analyses are based on retrospective and often single-centre series. No published or completed prospective cohort studies or randomized controlled trials were present in this literature search. We reviewed the case reports and cohort studies and where possible the local demographics. Because of the urgency to improve risk stratification in the middle of the ongoing pandemic, reports were considered that otherwise would not have met the rigors of a systematic review. All reports were assessed for risk of bias (ROB) using the Cochrane ROB 2.0 tool ^11^, however this assessment was used to inform the weighting given to the information contained therein when being reviewed by the experts in order to form a consensus risk assessment tool.

The nature of the risk tool was the subject of several focus group meetings. The requirement was for it to be simple to complete, be objective such that it could stratify vulnerability of exposure, and not reveal personal information such as may be misused by “line managers” after the pandemic. The latter requirement was a particular request of the Black, Asian and Minority Ethnic (BAME) representatives to the focus groups, who feel that they are particularly vulnerable to workplace bullying ^12^. As a result, the requirement for a single page risk assessment tool presenting cumulative factors that could be completed ahead of a conversation with the designated manager and present a clear stratification of vulnerability.

Risk of hospitalisation and mortality was analysed compared to population prevalence. Multivariate Cox regression modelling was used to estimate adjusted hazard ratio. Risk was normalised to a female aged 40-49, and an integer to approximate the impact of demographics, such as age^13^, ethnicity^14^ and important co-morbidities^15^ assigned.

There were two principle sources of data; the intensive care national audit and research centre (ICNARC) report which collated data from the national clinical audit covering all NHS adult, general intensive care and combined intensive care/high dependency units in England, Wales and Northern Ireland, plus some additional specialist and non-NHS critical care units, and the OPENSAFELY report which quantified a range of risk factors for death from COVID-19 based on primary care records^16^. Given these two principle sources of data, we collated and compared to the risk of admission to ITU and mortality from the ICNARC study with general population data ^17^. Predictive risk modelling was used to predict vulnerability of individuals.

This risk tool was standardised to the risk of mortality of a female under the age of 50 years. A point was then allocated for each approximate doubling in risk. Given the likely co-linearity of multiple risk factors where risk was a greater multiple than two it was rounded down. Since the purpose of this objective risk assessment tool is to supplement rather than supplant existing Public Health England recommendations, characteristics that warranted shielding according to the NHS Digital shielded patient list algorithm were discounted. Risk factors were only included in the derived objective risk assessment tool if they confidence interval of their independent predictive role did not cross the line of unity (i.e. p<0.05)

Once a simplified risk tool was compiled this was validated using the composite hazard ratios derived from the OpenSafely platform report^18^. We evaluated the risk in 317 cases within a trust and stratified them into low, middle and high risk. Agreement between the objective risk assessment tool and the calculated hazard ratio was evaluated using Cohen’s kappa coefficient for inter-rater agreement.

## Results

Multiple global observational studies were identified describing the risk of hospitalisation and mortality due to COVID-19, however there was significant heterogeneity in these studies, such that the robust nature of the data when applying to a UK population of health care providers was questionable (Supplementary table 1). One point of agreement, however, was that multiple co-morbidities appeared to confer cumulative risk. As a result, the development of a risk calculator was based exclusively on UK data, with multiple co-morbidities being given additive weighting.

### Clinician Demographics

#### Age and sex

In all age groups, mortality was at least twice as high in men as in women (Table 1). Compared to those under the age of 50, mortality was doubled in 50-59 year-olds, quadrupled in the 60-69 years age group, and 12 times higher after the age of 70 years in men.

**Table 1.**
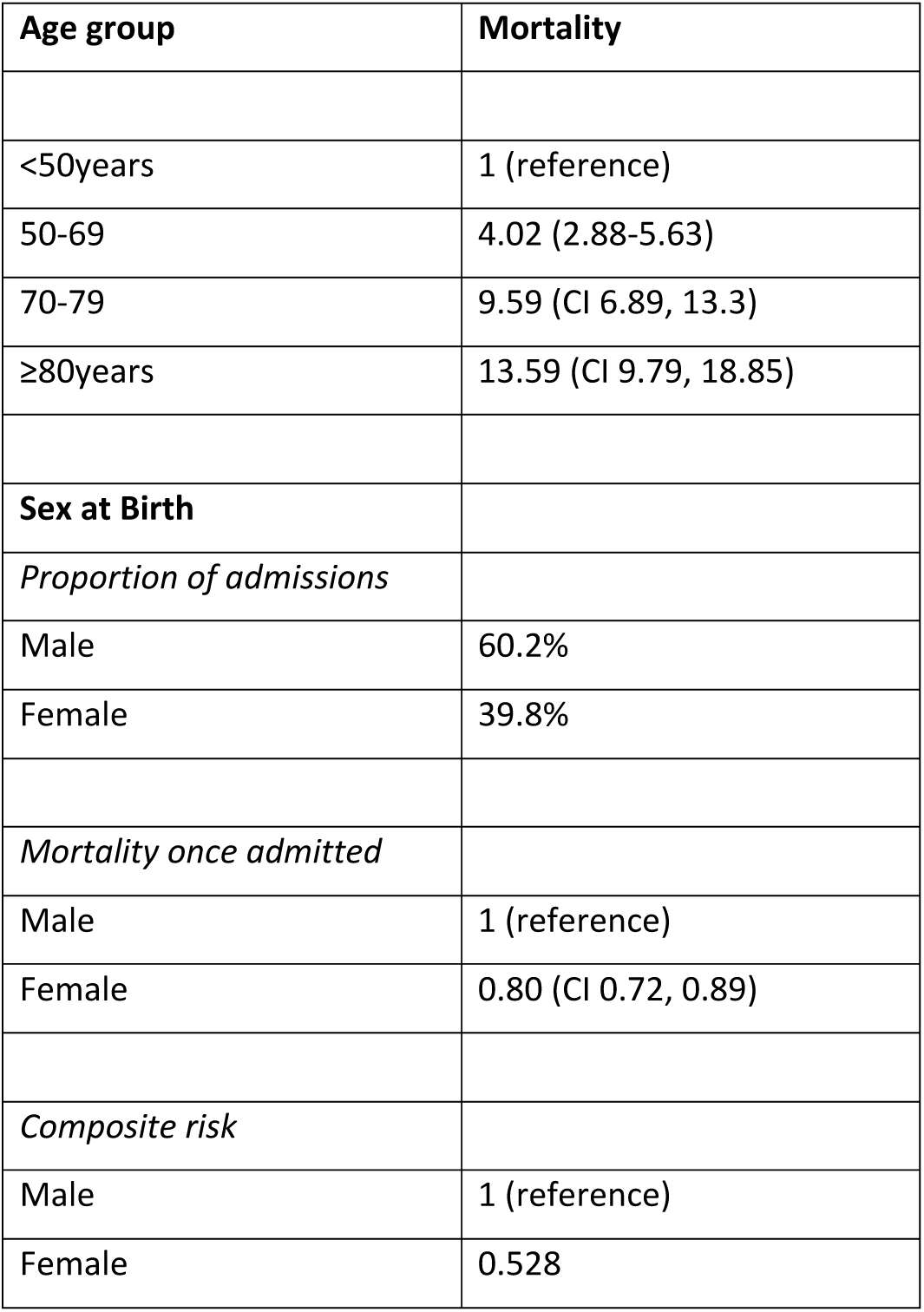
Clinician Demographics. Mortality by age group and risk of admission and subsequent mortality stratified by sex at birth (Features of 16,749 hospitalised UK patients with COVID-19 using the ISARIC WHO Clinical Characterisation Protocol) (18).

| Age group | Mortality |
| --- | --- |
| <50years | 1 (reference) |
| 50-69 | 4.02 (2.88-5.63) |
| 70-79 | 9.59 (CI 6.89, 13.3) |
| ≥80years | 13.59 (CI 9.79, 18.85) |
| **Sex at Birth** |  |
| *Proportion of admissions* |  |
| Male | 60.2% |
| Female | 39.8% |
| *Mortality once admitted* |  |
| Male | 1 (reference) |
| Female | 0.80 (CI 0.72, 0.89) |
| *Composite risk* |  |
| Male | 1 (reference) |
| Female | 0.528 |

### Ethnicity

People of non-white ethnic origin were at a higher risk of hospitalisation and mortality than the general population. Raw data suggested this was between 2-4 fold increased risk compared to the local population, and for COVID-19 compared to non-COVID-19 viral pneumoniae over the previous 3 years.^19^ This was, in part, explained by the premature onset of co-morbidities that also conferred risk, such as type 2 diabetes, ischaemic heart disease and stroke. ^20-23^ After multivariate adjustment, however, some of this risk could not be accounted for with conventional risk factors, and therefore an ethnicity adjustment was included. This is most accentuated in people of black African descent where the risk was two-fold elevated compared to those of white European descent. People of Indian Asian descent also had an approximately 50% increased risk of hospitalisation compared to their European counterparts.

### Socioeconomic status

As with flu, 25% of ICU admissions are people from the most deprived quintile as evaluated compared with just 15% from the least deprived (Table 2). Once on ITU, however, there were only slight differences in mortality between people in the most deprived vs least deprived status.

**Table 2.** Composite risk of contracting COVID-19 and mortality by pre-existing co-morbidity.

|  | Prevalence in<br>COVID-19<sup>19</sup> | Prevalence in<br>population | Relative risk of<br>contracting disease | Additional Risk of<br>mortality (18) | Composite<br>increased risk |
| --- | --- | --- | --- | --- | --- |
| Chronic Cardiac Disease | 29% | 14%<sup>33</sup> | 2.07 | 1.31 | 2.71 |
| Uncomplicated diabetes | 19% | 4.8%<sup>34</sup> | 3.95 | N/A | 3.95 |
| Chronic pulmonary disease<br>excluding asthma | 16% | 4.5%<sup>35</sup> | 3.56 | 1.19 | 4.2 |
| Asthma | 14% | 8.3%<sup>36</sup> | 2.15 | 1.19 | 2.55 |
| Dementia | 10% | 1.3%<sup>37</sup> | 7.69 | 1.39 | 6 |
| Malignant neoplasm | 9% | 1.5%<sup>38</sup> | 6.00 | 1.19 | 8.45 |
| On Immunosuppressant therapy | 9% | 0.8%<sup>39</sup> | 11.25 | N/A | 11.25 |
| Obesity (BMI>35kg/m<sup>2</sup>) | 38.5% | 27.8%<sup>34</sup> | 1.38 | 1.37 | 1.90 |
| Diabetes (with complications) | 6% | 1.2%<sup>34</sup> | 5.00 | N/A | 5 |

### Co-morbidities

There were multiple co-morbid factors that were each incrementally associated with increased mortality. The most common recorded comorbidities are chronic cardiac disease (29%), uncomplicated diabetes (19%), chronic pulmonary disease excluding asthma (19%) and asthma (14%) (Table 2) ^24^. These represented 16,749 patients: 7,924 (47%) patients had no documented reported comorbidity. Although numerically not a large percentage of patients, those with active malignant neoplasms, chronic kidney disease or liver disease were at between 3 and 5-fold increased risk of hospitalisation respectively compared to the prevalence in the general population. Although data was sparse, there was a suggestion that other conditions requiring long term immunosuppressant therapy was similarly over-represented by approximately 50% (data not shown). Similarly, dementia was associated with a significantly higher risk than the general population of both hospitalisation (∼7.7 times increased) and mortality in hospital (39% increase). This has limited relevance for modifying clinical exposure, although may be pertinent if using this tool to assess risk within the community. Contrary to many popular media reports, the increased risk of hospitalisation and mortality for people living with obesity was in the first stages accounted for by co-morbidities such as diabetes, ischaemic heart disease and stroke. Beyond a BMI of 35kg/m^2^ (or 30kg/m^2^ in people of Asian and Black African descent), however, there was an independent predictive increased risk.

### Generating an objective risk stratification tool

By considering each of the demonstrated associated factors for COVID-19 hospitalisation and subsequent mortality, a risk stratification tool was generated that may be considered when allocating clinical individuals to standard or higher risk duties (Table 3). The risk model attributes a point for every approximate doubling of risk compared to the reference population (Hazard Ratio ≥1.75 and ≤2.25). By adding the risk score from each category, it gives every individual a personal risk score which provides an estimate of their biological hazard.

**Table 3:** Suggested objective risk stratification (ORS) tool for individuals not already identified as “vulnerable” by the NHS Digital Shielded Patient List.

| Risk factor | Indicator | Adjustment |
| --- | --- | --- |
| Age | >50 | 1 |
|  | >60 | 2 |
|  | >70 | 4 |
|  | >80 | 6 |
| Sex at Birth | Female | 0 |
|  | Male | 1 |
| Ethnicity | Caucasian | 0 |
|  | Black African descent | 2 |
|  | Indian Asian descent | 1 |
|  | Filipino descent | 1 |
|  | Other (including Mixed race) | 1 |
| Diabetes and Obesity | (Type 1 or Type 2) uncomplicated* | 1 |
|  | (Type 1 or Type 2) complicated* | 2 |
| | BMI≥35kg/m<sup>2</sup> (>30kg/m<sup>2</sup> if BAME descent) | 1 |
| Cardiovascular disease | Angina, previous MI, stroke or cardiac intervention | 1 |
|  | Heart failure | 2 |
| Pulmonary disease | Asthma | 1 |
|  | Non-Asthma chronic pulmonary disease | 1 |
|  | Either above requiring oral corticosteroids in previous year | 1 |
| Malignant neoplasm | Active malignancy | 3 |
|  | Malignancy in remission | 1 |
| Chronic Kidney disease | CKD 3 or 4 | 2 |
|  | End stage renal disease/transplant | 4 |
| Chronic Liver disease | Any active disease | 3 |
| Immunosuppressant therapy | Any indication | 1 |
| **Interpretation** | **Score** |  |
| Low Risk | <3 |  |
| Medium Risk | 3-5 |  |
| High Risk | ≥6 | |

\*Complicated diabetes = presence of microvascular complications or HbA1c ≥ 64mmol/mol
CKD – Chronic Kidney disease

When validating this tool against the 317 predefined cases in a single NHS trust, the outcomes of the ORS tool correlated well with absolute risk scores in the OpenSafely platform (Cohen’s kappa 0.76 SD 0.071; p<0.0001; Table 4). A final validation was performed against the Public Health England document “Disparities in the risk and outcomes of COVID-19”.^10^ This demonstrated a similarly high level of agreement (Cohen’s kappa 0.81 SD 0.063; p<0.0001).

**Table 4:** Validation of the objective risk stratification tool compared to the OpenSafely Platform report. Number of healthcare workers scoring low, medium and high risk in a validation exercise of the two tools using data from 317 individuals working in the health care system.

| ORS tool \ | OpenSafely platform report |  |  | Total number of subjects |
| --- | --- | --- | --- | --- |
|  | ≤3 fold increased risk | 3-6 fold increased risk | ≥6 fold increased risk |  |
| Score <3 | 208 | 11 | 0 | 219 |
| PPV | 91.6% |  |  |  |
| Score 3-5 | 19 | 57 | 3 | 79 |
| PPV |  | 77.0% |  |  |
| Score ≥6 | 0 | 6 | 13 | 19 |
| PPV |  |  | 81.3% |  |
| Total | 227 | 74 | 16 | 317 |

ORS tool Objective Risk Stratification tool
(Cohen's kappa for agreement 0.76; p<0.0001)
PPV-positive predictive value

### Pregnancy

There is currently insufficient data to make any meaningful assessment about the risk of COVID-19 to either the mother or the unborn child. Early reports from the UK and the USA suggest there is no risk to either, however these are based on small numbers. ^25 26^ Given the unknown risk to both parties, although pregnancy is not considered as a risk factor in its own right, we would recommend all people who are pregnant be regarded as high risk and offered the option to shield.

## Discussion

There are currently no reliable data for COVID-19 related deaths in health care professionals including doctors; and surprisingly few data on the differences in risk in different healthcare settings. There is an urgent need for high quality research. We have applied general population risk factors to health care workers in order to generate a simplified biological risk stratification tool. This may serve to inform employers when allocating specific duties within the health care provision system, in order to fulfil their duty of care to their employees.

There are three types of risk for medical staff. The first relates to their biology, the second their environment and the third to the exposure. This tool evaluates the former in order to advise mitigation of the latter by stratifying individuals to lower, medium and higher risk. This biological risk assessment tool does not in any way replace the need for universal precautions with appropriate personal protective equipment. It should only be used to inform the need for modification of allocated duties to roles with little or no direct contact with patients, such as “advice and guidance” services, or virtual clinic provision. It incorporates and weights recognised risk factors. Many of these factors are predictable, such as age, gender, and pre-existing respiratory disease all of which have been associated with many previous viral infections such as H1N1 influenza.

The importance of pre-existing cardiovascular disease and cerebrovascular disease is a novel observation for a respiratory disease. This may be due to the method of cellular invasion of SARS-CoV-2 using the ACE2 enzyme; an enzyme which is responsible for physiological vascular health responses to hypertension and obesity. It does not, however, explain the risk associated with diabetes^27^, nor does it account for the increased risk in some ethnic groups.

A recent finding showed that Black, Asian and minority ethnic (BAME) individuals account for 63 per cent, 64 per cent and 95 per cent of deaths in the Nurse, Health Care Assistant and Doctor staff groups, respectively^1^. These figures are substantially higher than the proportional increase in BAME patients in UK intensive care units (mortality of 18% compared to 12% in the general population)^19^. Interestingly, our tool distinguished between people of Black African descent and people of other non-European backgrounds, awarding a higher risk to those of West African descent. When validating the ORS tool against the OpenSafely report, however, the differential point award demonstrated similar overall predictive role in people of Black African descent as other ethnicities. This is likely due to different confounding disease profile in these populations. People of Indian subcontinent heritage develop additional risk factors such as diabetes and premature cardiovascular disease approximately 10 years earlier than the European counterparts. People of Black African descent, however, are more likely to be affected by unmeasured risk factors such as haemoglobinopathies and systemic microvascular dysfunction ^28 29^.

### Application of the ORS tool

The primary role of any risk stratification tool is to provide a standardised approach to individual risk management by identifying those with the greatest hazard of adverse consequences from hazards.

Once individual risk is stratified, decisions regarding mitigating actions are required. Unfortunately, there remains uncertainty regarding the best action. The impact of recurrent exposure compared to high-risk exposures with high viral load, or the environment of the clinical domain is uncertain. Likewise, the relative impact of different environments has not adequately been assessed. Currently, employees in front-line emergency and acute medical settings such as A&E medicine, anaesthesia, respiratory medicine or gastroenterology may be considered at increased risk, as may be those who may need very close proximity with the patient such as ENT and ophthalmology. Some paradoxes have been observed. One recent paper found that the rate of infection with COVID-19 in staff in patient-facing occupations was no different from that in clerical/administrative staff without patient contact^30^ suggesting that PPE provides effective protection. Conversely, those later in the disease process with severe illness (particularly at the time of cytokine storm requiring high dependency care) may have reduced viral load and shedding^31^ therefore paradoxically have a lower potential to transmit infection compared to those at an early stage of the disease with no or relatively mild symptoms.

The ORS tool enables employers to decide when to exclude workers from working in presumed higher risk environments - even if workers do not wish to do so – or modify the nature of their duties, in order to fulfil the employers’ legal duty of care obligations to their work force. It must be acknowledged that this tool is based purely on biological risk of an individual. The prevalence of the disease in the community is another determinant which should be considered; when prevalence is low, the increased relative risk may not reflect a significant absolute risk, allowing health care practitioners to return to their usual role.

### Study limitations

Selection bias in testing, care and reporting can lead to differences in prevalence estimates of pre-existing risk factors and presentation across the reports from various countries. The majority of the existing analyses are based on retrospective and often single-centre series. No published or completed prospective cohort studies or randomised controlled trials were present in this literature search. A limitation is that we only searched Pubmed, EMBASE and preprint servers. There is an urgent need for high quality research, using individual level data for healthcare workers that will allow full mediation analyses in order to determine whether (for example) it is the age, the diabetes, or the cardiovascular disease that actually carries the greatest prognostic risk, given that these conditions commonly co-exist, and explore the disparity in BAME individuals between the general population and the healthcare deaths. There are currently only limited observational data for COVID-19 related deaths in health care workers or doctors, again without full access to all potentially pertinent information.. Importantly, this tool was derived from UK data, and therefore may not be relevant in other countries, however the methods employed here can be replicated in other healthcare settings.

### Patient and public involvement

The primary target of this research was healthcare professionals, occupational health teams and medical managers. There was significant engagement with members of the British Medical Association – the trade union representing UK doctors - COVID group, and the staff members. Several members of this group are listed as co-authors, including the chair of the representative body. It is important to distinguish that these individuals are reporting personal views based on their branch of practice and these are not necessarily the views of the Association.

### Concluding Remarks and Key Messages

As part of an employer’s legal obligation under the Health and Safety legislation all individuals are required to have a formal risk assessment. Although many organisations have advocated the need for individualised risk evaluation there remains no standardised methodology for this assessment. Without a consistent approach to stratification, comparing individuals’ personal risk within a department is difficult if not impossible. We have presented a robust scoring tool that allows comparisons and thus decisions to be made regarding the appropriate allocation of duties within a team. This also facilitates open discussion between staff who are being asked to work in patient facing areas and their team leaders, so they also understand their risks. All healthcare workers should wear appropriate PPE for any clinical examination or investigation on the basis that 20-40% of infected patients, especially if less than 40 years of age may be asymptomatic^32^. Within a specialty team, the highest risk individuals should be excluded from patient facing clinical areas; those at intermediate risk should have careful consideration to exclude them from front line areas or given limited duties avoiding close contact such as in ENT, ophthalmology and dentistry. Those at the lowest risk may be assigned duties with more patient contact. Neither the ORS tool, nor any other risk score, negates the need for good personal protective equipment and training.

## Data Availability

There is no new data - this represents an analysis of published data

## Contributor statement

JJ and WDS came up with the design and authored the first draft. WDS is responsible for the integrity of the analysis. All authors contributed to the format of the analysis and have contributed to the final manuscript.

## Data sharing statement

This manuscript is based on a secondary analysis of published data. The analysis plan and Stata output are available on contact with WDS

## Acknowledgements

The doctors from many branches of practice that gave comments and suggestions. We would also like to thank Professor Dame Parveen Kumar of Queen Mary University of London for helpful comments.

WDS is supported by the NIHR Exeter Clinical Research Facility and the NIHR Collaboration for Leadership in Applied Health Research and Care (CLAHRC) for the South West Peninsula.

The views expressed in this publication are those of the author(s) and not necessarily those of the NIHR Exeter Clinical Research Facility, the NHS, the British Medical Association, the NIHR or the Department of Health and Social Care in England.

## References

1. Cook T, Kursumovic E, Lennane S. Exclusive: deaths of NHS staff from covid-19 analysed: Wilmington Healthcare; 2020 [cited 2020 22 April]. Available from: Exclusive: deaths of NHS staff from covid-19 analysed Comment Health Service Journal accessed 1/5/20 2020.

2. Office for National Statistics O. Deaths involving COVID-19, England and Wales: deaths occurring in April 2020 2020 [Available from:https://www.ons.gov.uk/peoplepopulationandcommunity/birthsdeathsandmarriages/deaths/bulletins/deathsinvolvingcovid19englandandwales/deathsoccurringinapril2020 accessed 28 May 2020.

3. Givi B, Schiff BA, Chinn SB, et al. Safety Recommendations for Evaluation and Surgery of the Head and Neck During the COVID-19 Pandemic. JAMA Otolaryngol Head Neck Surg 2020 doi: 10.1001/jamaoto.2020.0780 [published Online First: 2020/04/02]

4. Greenland JR, Michelow MD, Wang L, et al. COVID-19 Infection: Implications for Perioperative and Critical Care Physicians. Anesthesiology 2020;132(6):1346–61. doi: 10.1097/ALN.0000000000003303 [published Online First: 2020/03/21]

5. Lai J, Ma S, Wang Y, et al. Factors Associated With Mental Health Outcomes Among Health Care Workers Exposed to Coronavirus Disease 2019. JAMA Netw Open 2020;3(3):e203976. doi: 10.1001/jamanetworkopen.2020.3976[published Online First: 2020/03/24]

6. Lai THT, Tang EWH, Chau SKY, et al. Stepping up infection control measures in ophthalmology during the novel coronavirus outbreak: an experience from Hong Kong. Graefes Arch Clin Exp Ophthalmol 2020;258(5):1049–55. doi: 10.1007/s00417-020-04641-8 [published Online First: 2020/03/04]

7. NHS England. Patient Deaths report 2020 [Available from: https://www.england.nhs.uk/statistics/statistical-work-areas/covid-19-daily-deaths/ accessed 1st May 2020.

8. NHS Employers. Risk assessments for staff 2020 [Available from: https://www.nhsemployers.org/covid19/health-safety-and-wellbeing/risk-assessments-for-staff accessed 29th May 2020.

9. Khunti K, Singh AK, Pareek M, et al. Is ethnicity linked to incidence or outcomes of covid-19? BMJ 2020;369:m1548. doi: 10.1136/bmj.m1548 [published Online First: 2020/04/22]

10. Public Health England. Disparities in the risk and outcomes of COVID-19 2020 [Available from: https://assets.publishing.service.gov.uk/government/uploads/system/uploads/attachment_data/file/889195/disparities_review.pdf accessed 2nd June 2020.

11. Higgins JPT, Altman DG, Gøtzsche PC, et al. The Cochrane Collaboration’s tool for assessing risk of bias in randomised trials. BMJ 2011;343:d5928. doi: 10.1136/bmj.d5928

12. Carter M, Thompson N, Crampton P, et al. Workplace bullying in the UK NHS: a questionnaire and interview study on prevalence, impact and barriers to reporting. BMJ Open 2013;3(6):e002628. doi: 10.1136/bmjopen-2013-002628

13. Verity R, Okell LC, Dorigatti I, et al. Estimates of the severity of coronavirus disease 2019: a model-based analysis. Lancet Infect Dis 2020 doi: 10.1016/S1473-3099(20)30243-7 [published Online First: 2020/04/03]

14. Office of National Statistics. UK population by ethnicity 2020 [Available from: https://www.ethnicity-facts-figures.service.gov.uk/uk-population-by-ethnicity accessed 1st May 2020.

15. Wang B, Li R, Lu Z, et al. Does comorbidity increase the risk of patients with COVID-19: evidence from meta-analysis. Aging (Albany NY) 2020;12(7):6049–57. doi: 10.18632/aging.103000 [published Online First: 2020/04/09]

16. Williamson E, Walker AJ, Bhaskaran KJ, et al. OpenSAFELY: factors associated with COVID-19-related hospital death in the linked electronic health records of 17 million adult NHS patients. medRxiv 2020:2020.05.06.20092999. doi: 10.1101/2020.05.06.20092999

17. Doidge JC, Gould DW, Ferrando-Vivas P, et al. Trends in Intensive Care for Patients with COVID-19 in England, Wales and Northern Ireland. Am J Respir Crit Care Med 2020 doi: 10.1164/rccm.202008-3212OC [published Online First: 2020/12/12]

18. Coggon D, Croft P, Cullinan P, et al. Assessment of Workers’ Personal Vulnerability to COVID-19 Using “COVID-AGE”. medRxiv 2020:2020.05.21.20108969. doi: 10.1101/2020.05.21.20108969

19. Docherty AB, Harrison EM, Green CA, et al. Features of 16,749 hospitalised UK patients with COVID-19 using the ISARIC WHO Clinical Characterisation Protocol. medRxiv 2020:2020.04.23.20076042. doi: 10.1101/2020.04.23.20076042

20. Hills AP, Arena R, Khunti K, et al. Epidemiology and determinants of type 2 diabetes in south Asia. Lancet Diabetes Endocrinol 2018;6(12):966–78. doi: 10.1016/s2213-8587(18)30204-3 [published Online First: 2018/10/06]

21. Bello O, Mohandas C, Shojee-Moradie F, et al. Black African men with early type 2 diabetes have similar muscle, liver and adipose tissue insulin sensitivity to white European men despite lower visceral fat. Diabetologia 2019;62(5):835–44. doi: 10.1007/s00125-019-4820-6 [published Online First: 2019/02/08]

22. Smeeton NC, Heuschmann PU, Rudd AG, et al. Incidence of hemorrhagic stroke in black Caribbean, black African, and white populations: the South London stroke register, 1995–2004. Stroke 2007;38(12):3133–8. doi: 10.1161/strokeaha.107.487082 [published Online First: 2007/10/27]

23. Wolfe CD, Rudd AG, Howard R, et al. Incidence and case fatality rates of stroke subtypes in a multiethnic population: the South London Stroke Register. J Neurol Neurosurg Psychiatry 2002;72(2):211–6.

24. Wynants L, Van Calster B, Bonten MMJ, et al. Prediction models for diagnosis and prognosis of covid-19 infection: systematic review and critical appraisal. BMJ 2020;369:m1328. doi: 10.1136/bmj.m1328 [published Online First: 2020/04/09]

25. Knight M, Bunch K, Vousden N, et al. Characteristics and outcomes of pregnant women admitted to hospital with confirmed SARS-CoV-2 infection in UK: national population based cohort study. BMJ 2020;369:m2107. doi: 10.1136/bmj.m2107

26. Adhikari EH, Moreno W, Zofkie AC, et al. Pregnancy Outcomes Among Women With and Without Severe Acute Respiratory Syndrome Coronavirus 2 Infection. JAMA Netw Open 2020;3(11):e2029256. doi: 10.1001/jamanetworkopen.2020.29256 [published Online First: 2020/11/20]

27. Tikellis C, Johnston CI, Forbes JM, et al. Characterization of renal angiotensin-converting enzyme 2 in diabetic nephropathy. Hypertension 2003;41(3):392–7. doi: 10.1161/01.HYP.0000060689.38912.CB [published Online First: 2003/03/08]

28. Strain WD, Chaturvedi N, Leggetter S, et al. Ethnic differences in skin microvascular function and their relation to cardiac target-organ damage. J Hypertens 2005;23(1):133–40. doi: 00004872-200501000-00023 [pii] [published Online First: 2005/01/12]

29. Strain WD, Chaturvedi N, Nihoyannopoulos P, et al. Differences in the association between type 2 diabetes and impaired microvascular function among Europeans and African Caribbeans. Diabetologia 2005;48(11):2269–77. doi: 10.1007/s00125-005-1950-9 [published Online First: 2005/09/30]

30. Hunter E, Price DA, Murphy E, et al. First experience of COVID-19 screening of health-care workers in England. Lancet 2020;395(10234):e77–e78. doi: 10.1016/S0140-6736(20)30970-3 [published Online First: 2020/04/26]

31. Wölfel R, Corman VM, Guggemos W, et al. Virological assessment of hospitalized patients with COVID-2019. Nature 2020;581(7809):465–69. doi: 10.1038/s41586-020-2196-x

32. Zhou P, Yang XL, Wang XG, et al. A pneumonia outbreak associated with a new coronavirus of probable bat origin. Nature 2020;579(7798):270–73. doi: 10.1038/s41586-020-2012-7 [published Online First: 2020/02/06]

33. NHS Digital. Health Survey for England 2017 2018 [Available from: Health Survey for England 2017 [NS] - NHS Digital accessed 1st May 2020.

34. Diabetes UK. Facts and figures 2019 [Available from: https://www.diabetes.org.uk/professionals/position-statements-reports/statistics accessed 28th April 2020.

35. British Lung Foundation. The UK big picture 2019 [Available from: https://statistics.blf.org.uk/lung-disease-uk-big-picture accessed 28th April 2020.

36. Asthma UK. Facts and Figures 2019 [Available from: https://www.asthma.org.uk/about/media/facts-and-statistics/ accessed 28th April 2020.

37. Alzheimers UK. Dementia Statistics Hub 2018 [Available from: https://www.dementiastatistics.org/statistics-about-dementia/prevalence/ accessed 28th April 2020.

38. Cancer Research UK. Cancer Statistics for the UK 2019 [Available from: https://www.cancerresearchuk.org/health-professional/cancer-statistics-for-the-uk accessed 28th April 2020.

39. Arthritis UK. The state of musculoskeletal health 2019 2019 [Available from: https://www.versusarthritis.org/media/14594/state-of-musculoskeletal-health-2019.pdf accessed 28th April 2020.

